# A data-driven method to detect the flattening of the COVID-19 pandemic curve and estimating its ending life-cycle using only the time-series of new cases per day

**DOI:** 10.1101/2020.05.15.20103374

**Authors:** P. Gupta, K. K. Sharma, S.D. Joshi, S. Goyal

## Abstract

The novel Coronavirus-19 disease (COVID-19) has emerged as a pandemic and has presented itself as an unprecedented challenge to the majority of countries worldwide. The containment measures for this disease such as the requirement of health care facilities greatly rely on estimating the future dynamics and flattening of the COVID-19 curve. However, it is always challenging to estimate the future trends and flattening of the COVID-19 curve due to the involvement of many real-life variables. Recently, traditional methods based on SIR and SEIR have been presented for predictive monitoring and detection of flattening of the COVID-19 curve. In this paper, a novel method for detection of flattening of the COVID-19 curve and its ending life-cycle using only the time-series of new cases per day is presented. Simulation results are compared to the SIR based methods in three different scenarios using COVID-19 curves for South Korea, the United States of America, and India. In this study, simulations, performed on the 26^th^ April 2020 show that the peak of the COVID-19 curve in the USA has already arrived and situated on the 14^th^ of April 2020, while the peak of the COVID-19 curve for India has yet to arrive.

## 1. Introduction

The novel coronavirus was first found in the Chinese city of Wuhan, in December 2019 [1–3]. Soon the European countries were also affected by this coronavirus and there was a rapid rise in new cases per day, which in turn led to increasing the accumulated cases [4, 5]. Due to the absence of a vaccine and serious implications of the acute respiratory syndrome (ARS) this situation has presented itself as an unprecedented challenge to the whole of the world specifically to western Europe and the USA.

The common characteristic of the COVID-19 spread across the nations is that there is an increase in the new cases per day initially, then flattens and comes to an end after following almost an exponential trend. The graphical representation of new cases per day is known as the COVID-19 curve. At this stage, everyone wants to know when the curve will flatten, and when this will end. This is important to note that the prolonged flattening of the curve may not only put an extra burden on the healthcare system but also to the economic activities of a nation.

Thus, it is always a challenge for those who are involved in policy interventions and assessing health care requirements to estimate the present trends and the possibility of flattening of the COVID-19 curve shortly. This assessment ultimately helps to make suitable decisions in the future. Traditionally, estimation of epidemiological disease dynamics is carried out using susceptible-infected-recovered (SIR) and susceptible-infected-exposed-recovered (SIER) based models. Recently, an effort has been made to detect flattening and estimating the ending life-cycle of COVID-19 curves based on the SIR model [6]. However, it is reported that the main disadvantage of SIR based methods is that they require estimates of critical epidemiological parameters [7].

In this paper, a novel method for the detection of the flattening of the COVID-19 curve and estimating the ending life-cycle is presented, which require only the time-series of new cases per day (NCPD) and doesn’t require critical epidemiological parameters

## 2. Proposed method

The proposed method and the basic monitoring mechanism are explained through the block diagram as shown in Fig. 1. In the proposed method, first, a time-series NCPD is formed. The data points of the NCPD time-series is then used to capture its average behaviour using a Gaussian function represented by the following equation:

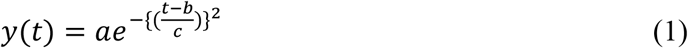

**Figure 1:**
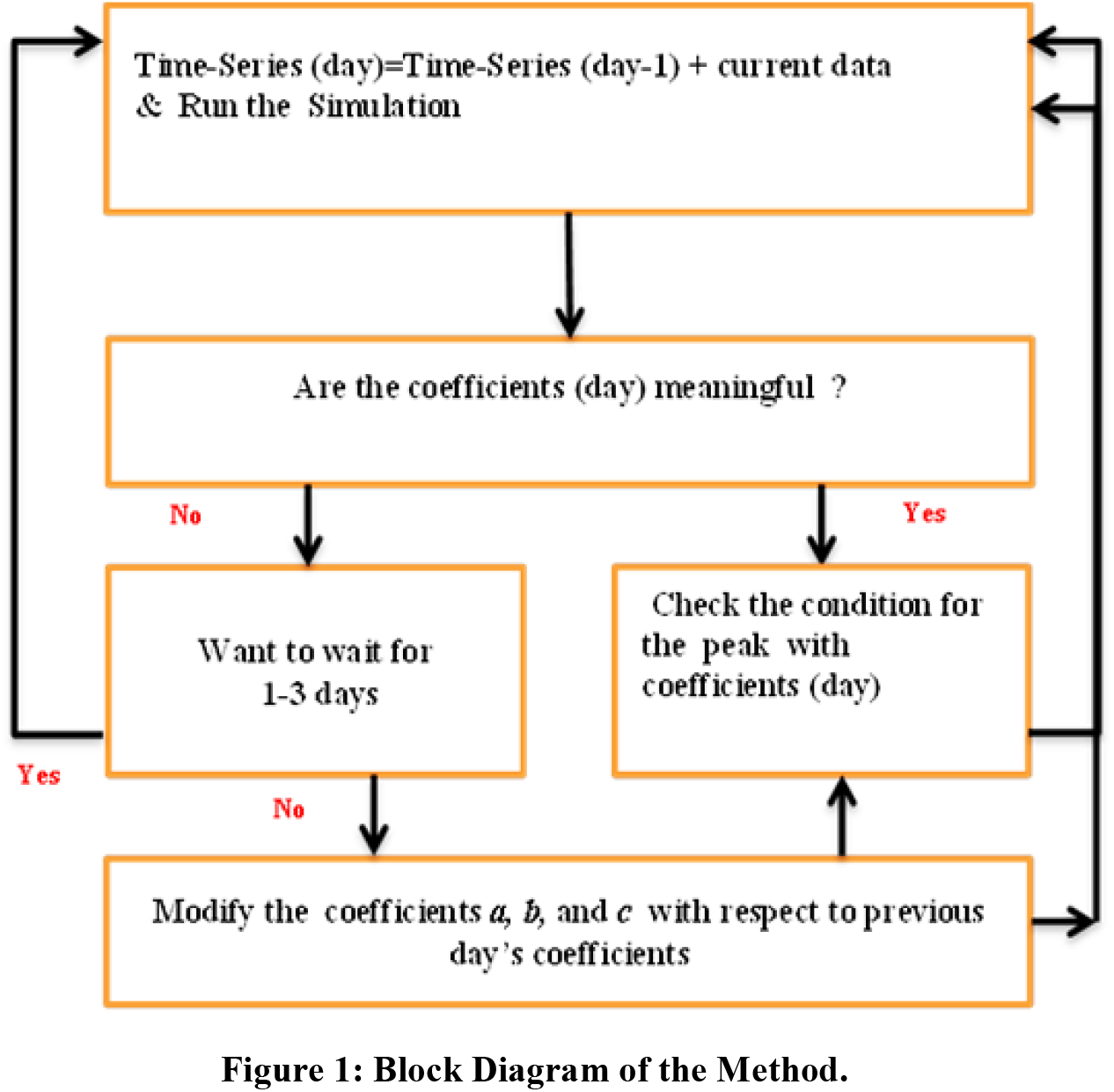
Block Diagram of the Method.

here, *t* is the time in days, and *y(t)* is the estimate of the average behaviour of the NCPD time-series. We are considering only positive values of *t*. Coefficients *a, b* and *c* represent the amplitude of the peak, centroid (location),and width of the curve respectively, representing average behaviour of the NCPD time-series. Coefficients of this equation are obtained after fitting the average behaviour of this NCPD time-series to the mentioned Gaussian function. In our method, this equation is chosen because of two reasons. First, that we are mainly concerned with the major portion of the COVID-19 curve, which contributes to the first wave such as the first wave of the COVID-19 curve of China. Second, the flattening of the curve is usually expected in the first half of the major portion of the curve. After obtaining the coefficients *a, b* and *c* of the eqn. (1), estimation of the peak is made and the relative position of the peak is also obtained. A positive value of the relative peak position indicates, how far the peak is from the current day, while the negative value indicates that peak has already arrived.

In the event when the current value of the data point greatly differs from the previous values of the data points, it is proposed to wait for 1–3 days until spread dynamics stabilizes. Even if, the future data points are to be estimated on these days, the current data value is iterated and modified until the meaningful coefficients are obtained.

Here, meaningful coefficients represent those values of a*, b*, and *c* which allow the spread dynamics to attain realizable values shortly. For example, the relative peak position abruptly changes on the current day from 5 days to 70 days is not realizable in the near future, however, a change in peak position from 5 to 25 days is realizable. In this manner, using eqn. (1), ending the life-cycle of the curve is also estimated. Here, ending life-cycle means the day when the curve will come to an end. For example, the ending date of 97% life-cycle of the curve is a day when 97% of the total expected cases will be reported.

## 3. Simulation results and discussion

We simulated, the NCPD time-series for three different countries i.e., South Korea, the USA, and India. To test our methodology, first, we compared our simulation results to the already known results of South-Korea (Here the major COVID-19 curve has already ended). Then, we compared our simulation results to the SIR based method for the detection of flattening and estimating the ending life-cycle of COVID-19 curves [6]. The results of this method [6], for some of the countries, were simulated on the 26^th^ of April 2020 and have been published in the pdf format on the website. We also used a similar data source [8], except the data for India on the 9^th^ of March 2020, which was missing. For the missing data, we have used the data from the WHO website [9]. All simulations consider the first day as the day when the first laboratory tested positive case was reported and counting of days is done from this first date. We divided our simulation results into three scenarios as explained below:

### First Scenario-The COVID-19 curve of South-Korea

The peak of the COVID-19 curve has arrived in South-Korea and the major portion of the curve has already ended. The COVID-19 curve for South-Korea is a good example to explain the dynamics of the peak arrival and not changing its relative position.

The first case in South-Korea was reported on the 20^th^ of January 2020. Results of our simulations, dated 3^rd^ of March 2020 (44^th^ day) show that the peak has arrived and has a position on the 2^nd^ of March 2020. Further simulations, on consecutive days confirmed that the peak position is almost constant. This can be observed in Fig. 2, Fig. 3 (a), and Fig. 3 (b). Figure 2, confirms the peak arrival, as the current position (in green colour) is right to the peak position (in red colour)

**Figure 2:**
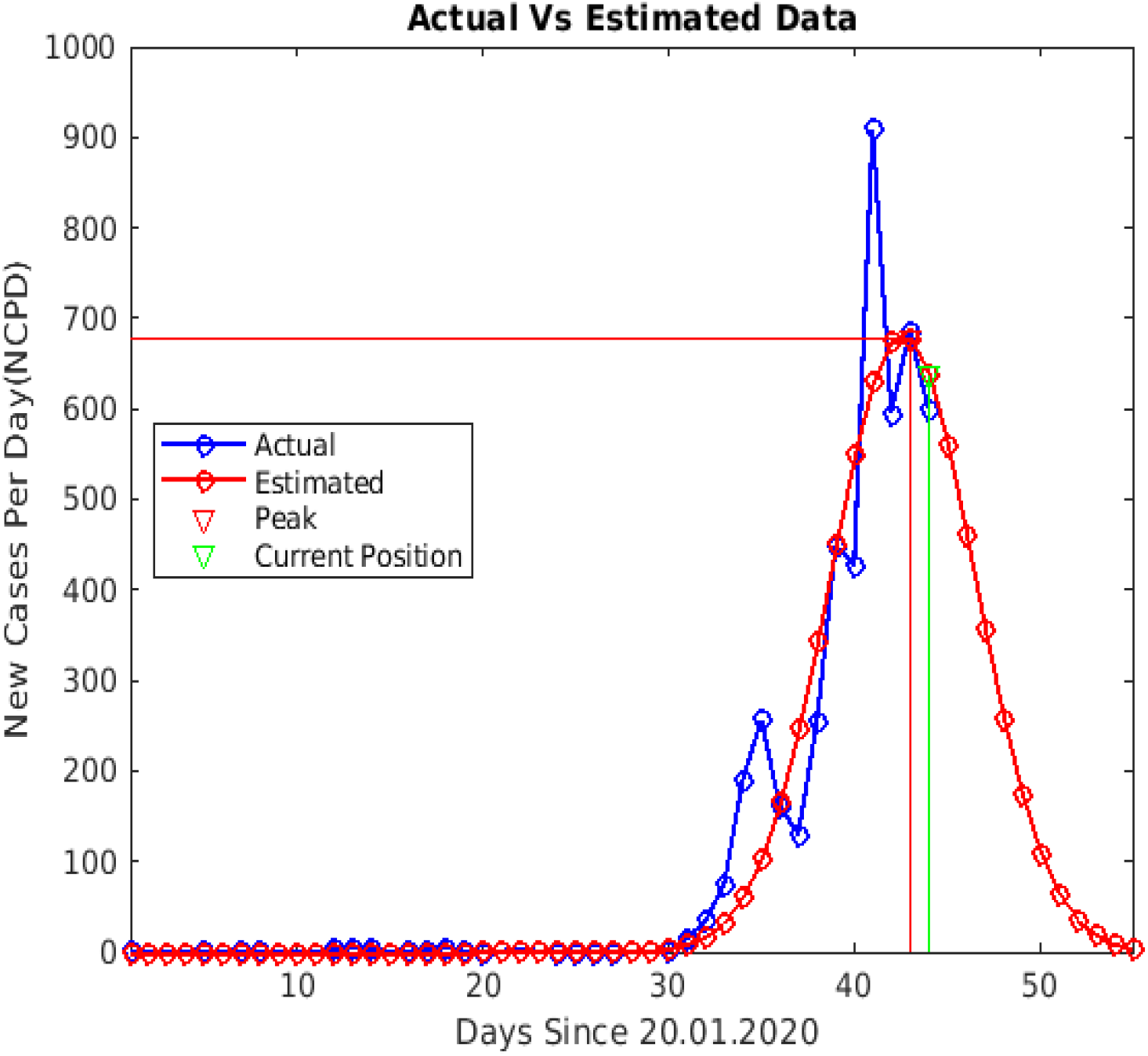
Simulation on the 44^th^ day (green) for South-Korea confirms the peak arrival on the 43^rd^ day (red) and estimation of possible ending.

**Figure 3:**
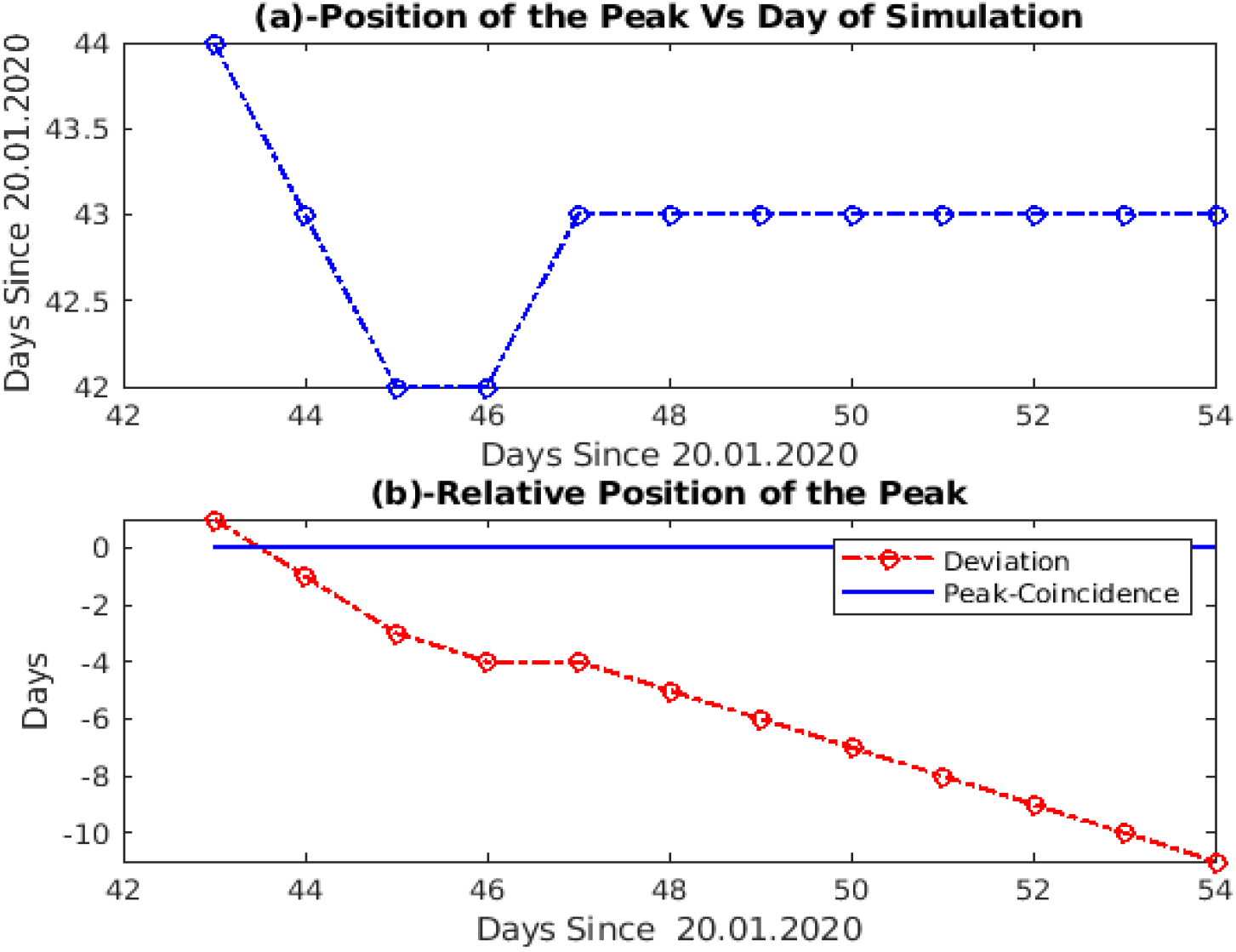
(a) Simulations from the 43^rd^ day to the 54^th^ day show that the peak position is almost constant (b) relative position of the peak is negative and decreasing fast.

Figure 3(a) indicates the peak positions on the day of simulations. For example simulation on 44^th^ day shows peak position on 43^rd^ day, which generate a negative value of one day for the relative position of the peak. This is indicated in Fig. 3(b). This can also be seen in Fig. 3(a), that the position of the peak to the progressing days of simulations remains almost constant. This situation indicates an ideal condition for falling of the COVID-19 curve.

Figure 4, shows simulation results of the 54^th^ day and indicates almost constant peak value as compared to that on the 44^th^ day (Fig.2). Figure 4, also shows that the estimated data points grossly fit the actual data points to the end of the curve. One can also see the difference in curve dynamics between the 44^th^ day and the 54th day in Fig. 2 and Fig. 5 respectively. The future data points, which were estimated by the method on the 44^th^ day were almost followed until the 53^rd^ day.

**Figure 4:**
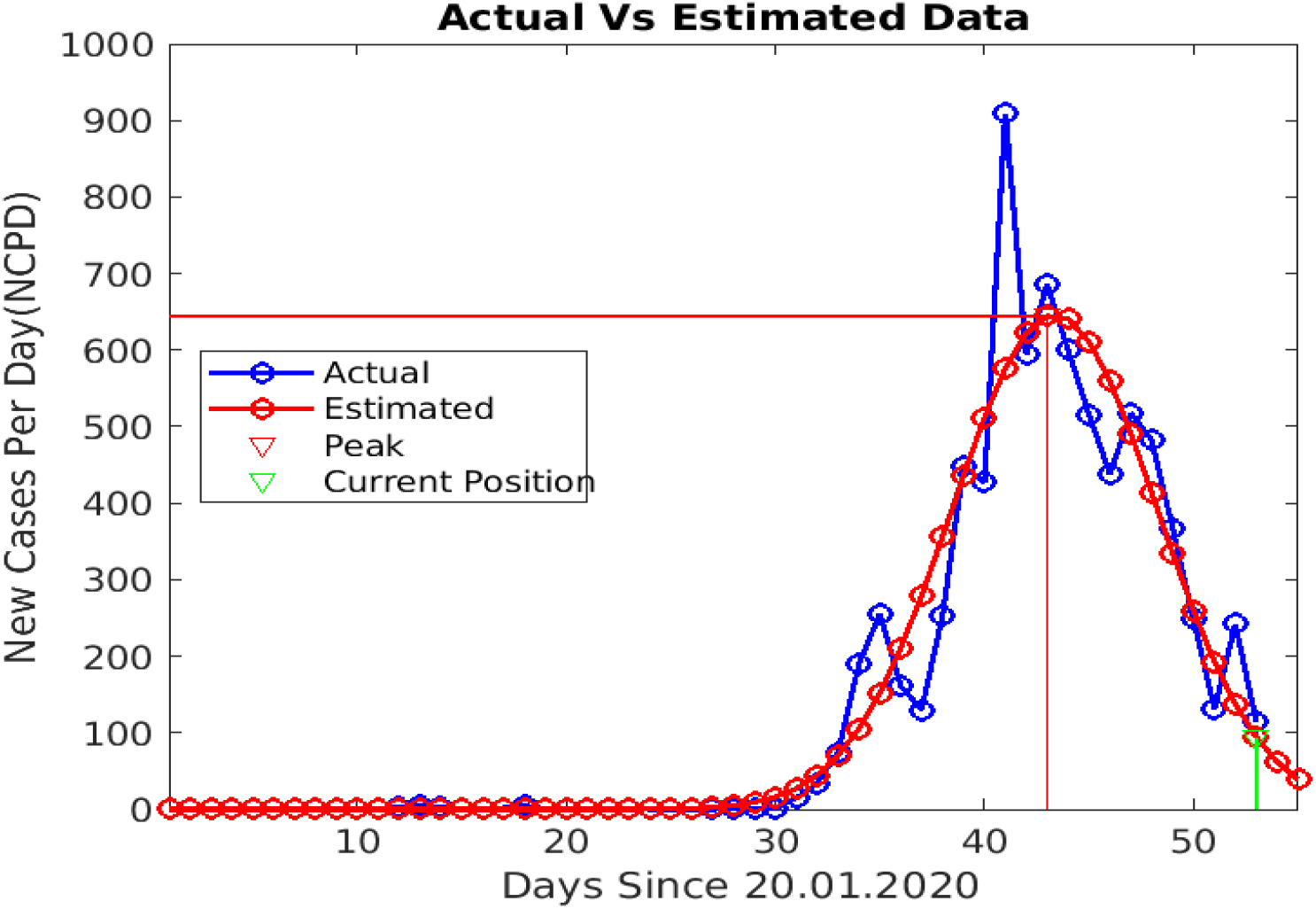
Simulation on the 54^th^day for South-Korea shows that peak amplitude is almost constant as compared to the 44^th^ day (Fig.2) and a major portion of the curve is grossly fitted to the actual data points.

**Figure 5:**
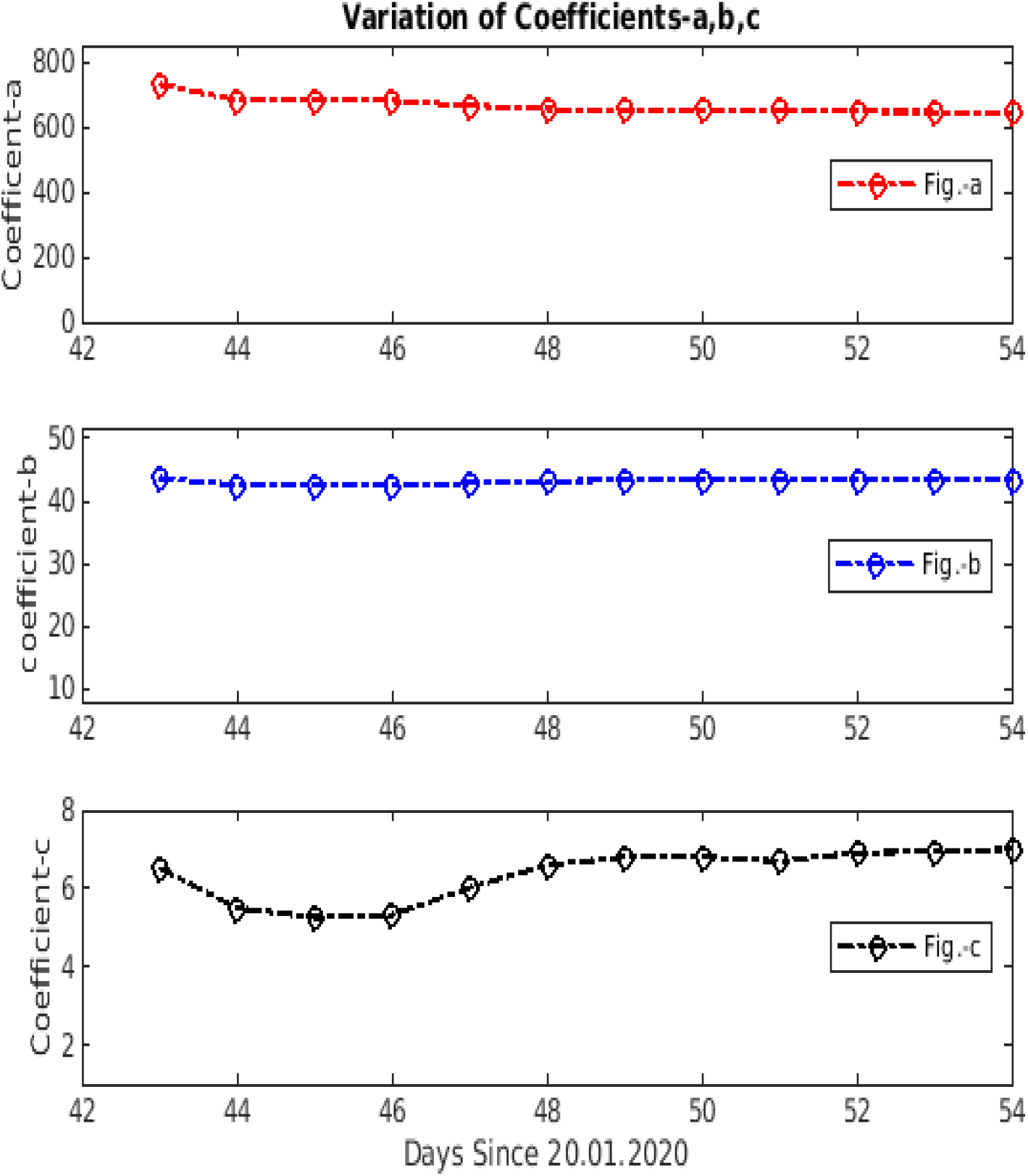
(a) Variation in the value of coefficient-*a* from the 43^rd^ day to the 54th day (b) variation in the value of coefficient-*b* (c) variation in the value of coefficient-*c*.

We also investigated the variation in values of coefficients *a, b* and *c*. Figure 4(a), 4(b), and 4(c) indicates that the these values don’t change significantly from the 44^th^ day to the 54^th^ day, which shows that the dynamics of the South-Korea curve got stabilized

### Second Scenario-The COVID-19 curve of the USA

The peak of the COVID-19 curve has arrived in the USA but the COVID curve is not yet ended. Also, the peak is continuously changing its relative position with negative values.

The first case in the USA was reported on the 21^st^ of January 2020. Our simulations, performed on dated 26^th^ April 2020, show that the peak is situated on the 85^th^ day (14^th^ April). This can be seen in Fig. 6(a). Figure 6(a) and 6(b) also indicates that the peak is continuously changing its relative position as contrary to the previous scenario of South-Korea. This explains the extended life-cycle for the COVID-19 curve of the USA, despite there was a peak on the 77^th^ day.

**Figure 6:**
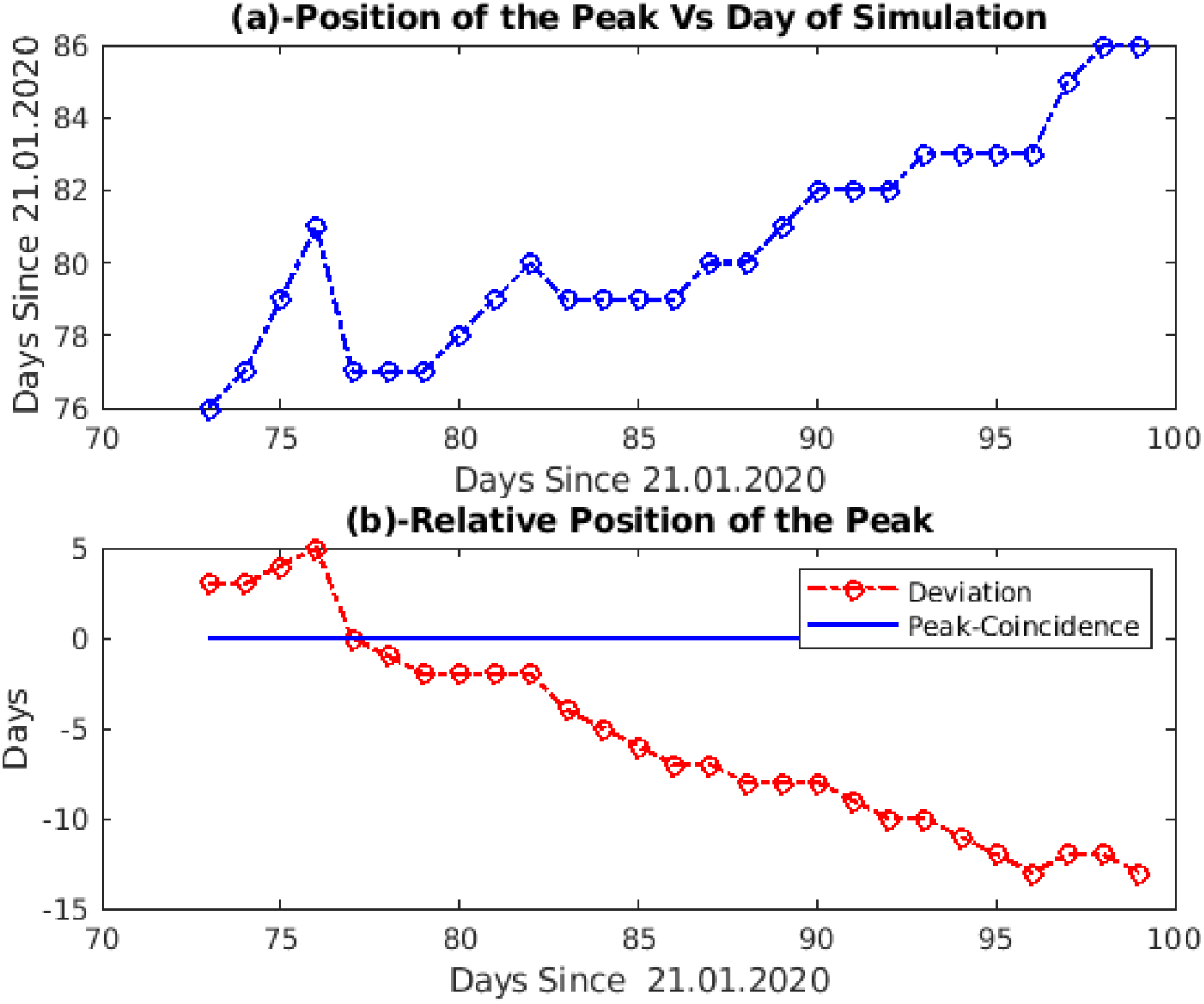
(a) Simulations from the 73^rd^ day to the 99^th^ day show that the peak position is varying (b) relative positions of the peak is negative but not decreasing fast.

The simulation results on the 99^th^ day also explain the changing dynamics of the COVID-19 curve between the 78^th^ and the 99^th^ day as shown in Fig. 7.

**Figure 7:**
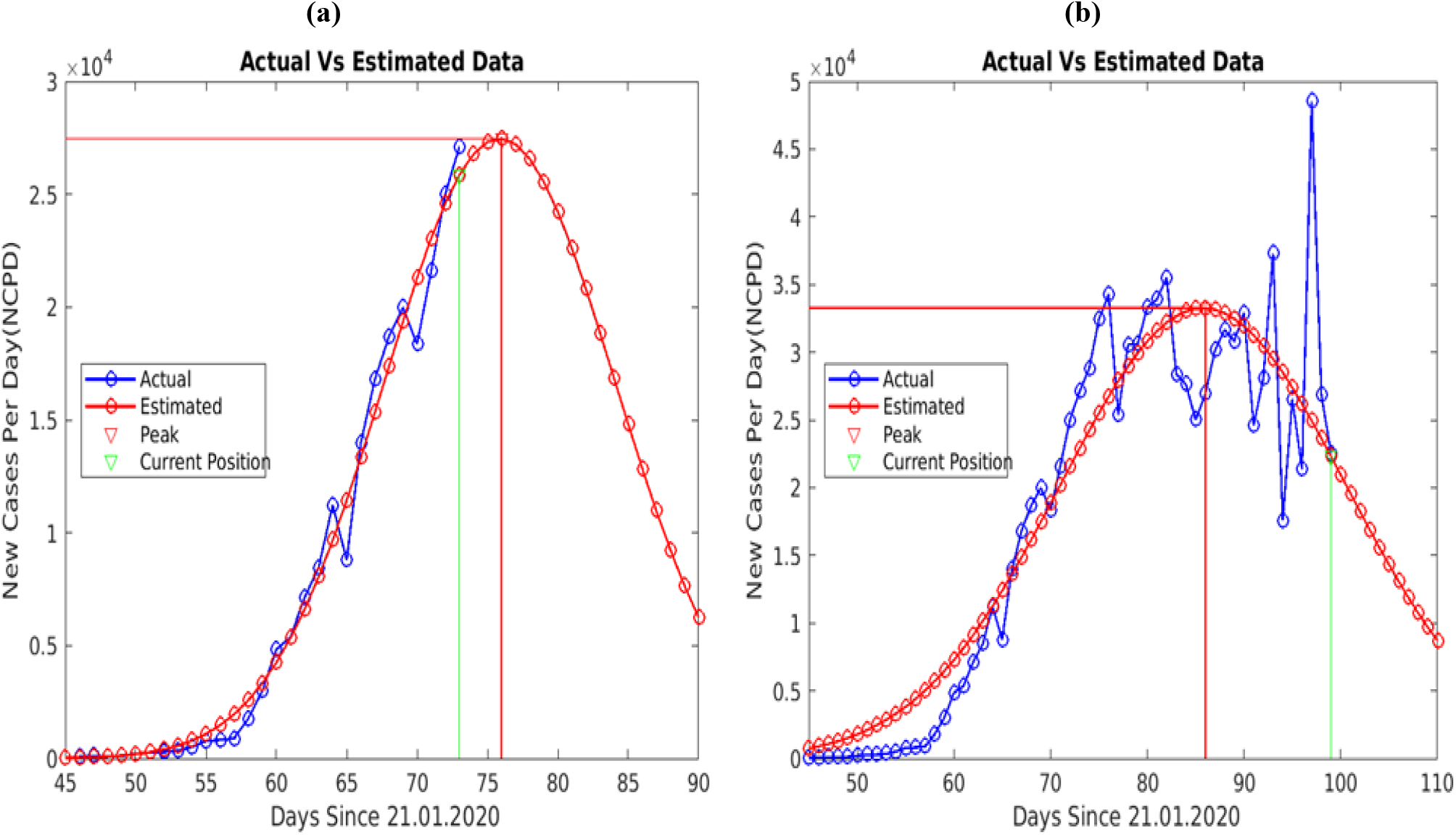
(a) Simulation on the 78^th^ day (green) shows the amplitude of the peak, its position (red) and, fitting of actual data points (b) simulation on the 99^th^ day in the USA shows that the peak amplitude has increases as compared to peak value on 78^th^ day and also changed its position from the 77^th^ day to the 86^th^ day. This will result in delayed ending and more accumulated cases.

The investigations on the variation in values of coefficients *a, b*, and *c* as shown in Fig. 8, also indicate the changing dynamics of the USA curve from the 73^rd^ day to the 99^th^ day.

**Figure 8:**
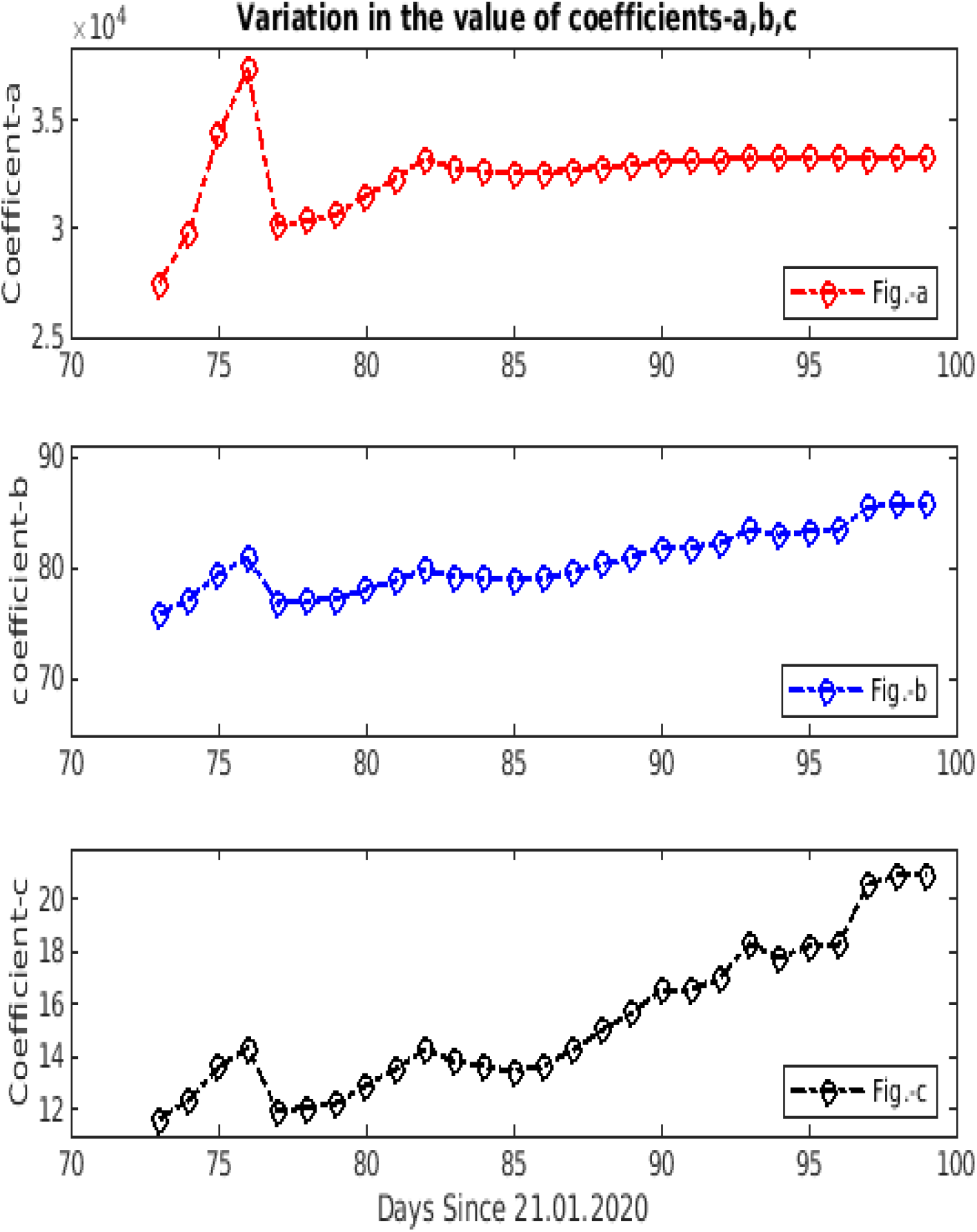
(a) Variation in the value of coefficient-*a* from the 73^rd^ day to the 99^th^ day (b) variation in the value of coefficient-*b* (c) variation in the value of coefficient-*c*

### Third Scenario-The COVID-19 curves for India

The peak of the COVID-19 curve has not been observed on the day of simulation (26th April 2020).

The first case in India was reported on the 30^th^ of January 2020. Our simulations, performed on dated 26^th^ April 2020, show that the peak is situated on the 95^th^ day (3^rd^ May 2020). Figure 9 (a) indicates peak position on simulation days, while Fig. 9 (b) represents its relative values on those days. Figure 10 (b), also indicates that a peak was reported to arrive on 79^th^ day (17th April) but this peak was in a transient phase and came out from this phase on the 81^st^ day (19th April) and still struggling to fall back to the relative negative peak position.

**Figure 10:**
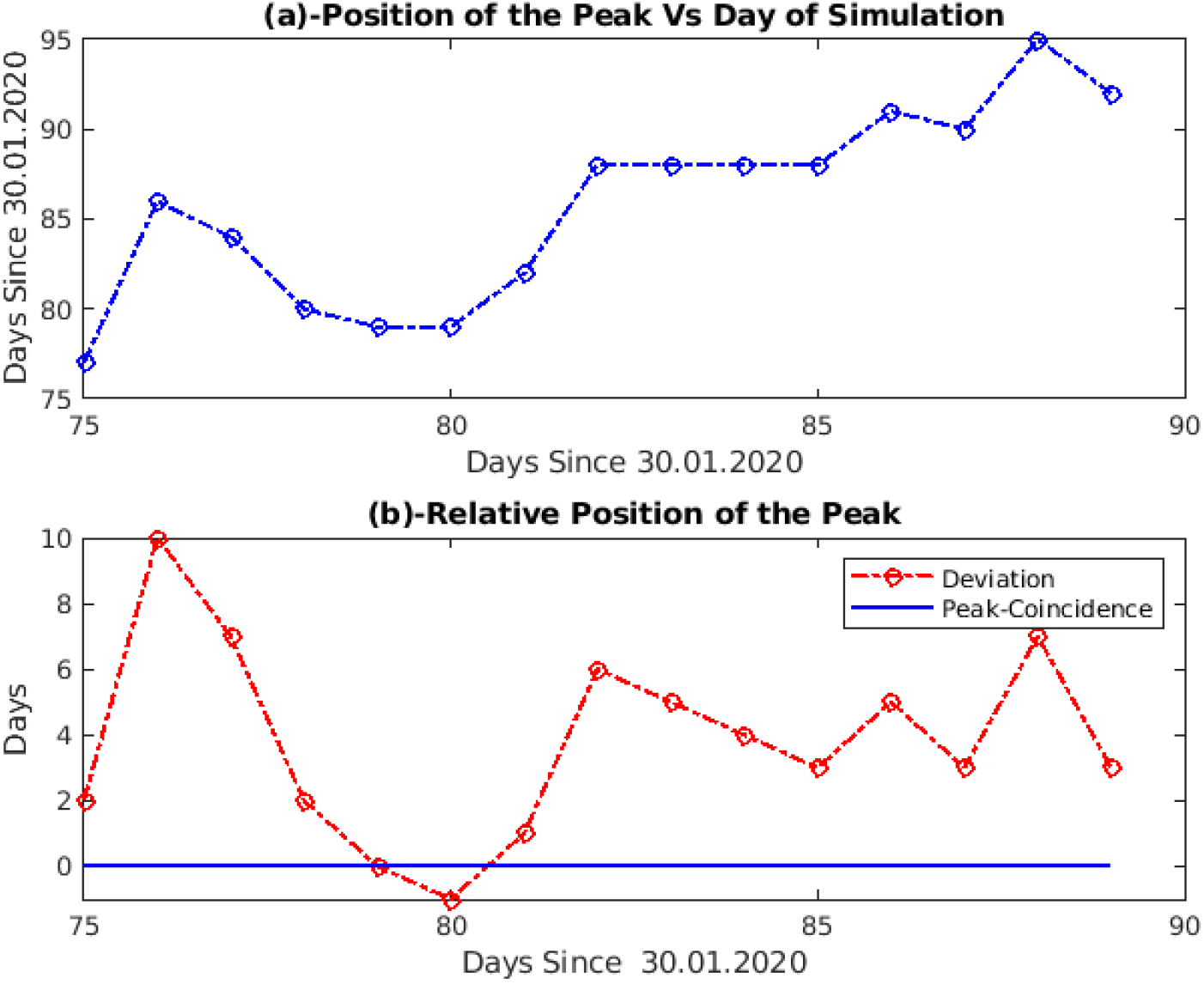
(a) Simulations from the 75^th^ day to the 89^th^ day show that the peak position is varying (b) relative positions of the peak is positive except on the 80^th^ day and coinciding on the 79^th^ day

Simulation results for India, for the 80^th^ and 89^th^ day, shown in Fig. 11, also indicate the change in the peak amplitude of the COVID-19 curve, which will not only result in more cumulative cases but also the prolonged ending life-cycle.

**Figure 11:**
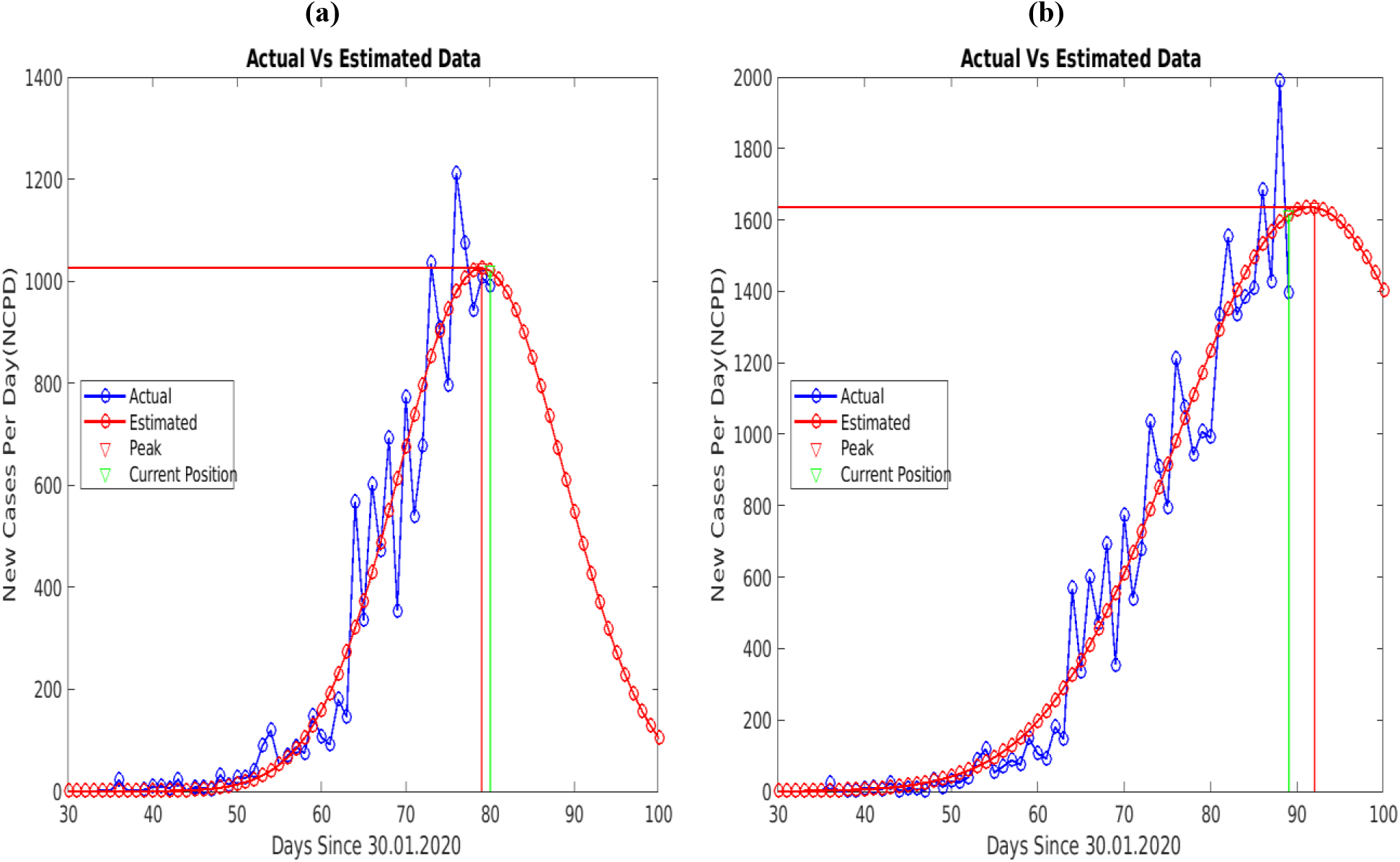
(a) Simulation on the 80^th^ day (green) shows the amplitude of the peak, its position (red) and the actual data points. (b) Simulation results on the 89^th^ day (green) show that the amplitude of the peak has increased which will result in delayed ending and more accumulated cases in India.

We also simulated a possible ending of these COVID-19 curves for the USA and India on dated 26^th^ of April 2020, while for South–Korea, we simulated this situation before it was ended. The comparisons of results for peak position and estimates of 97% and 99% ending of the curve are tabulated in Table-1 and Table-2 respectively

**Table 1:**
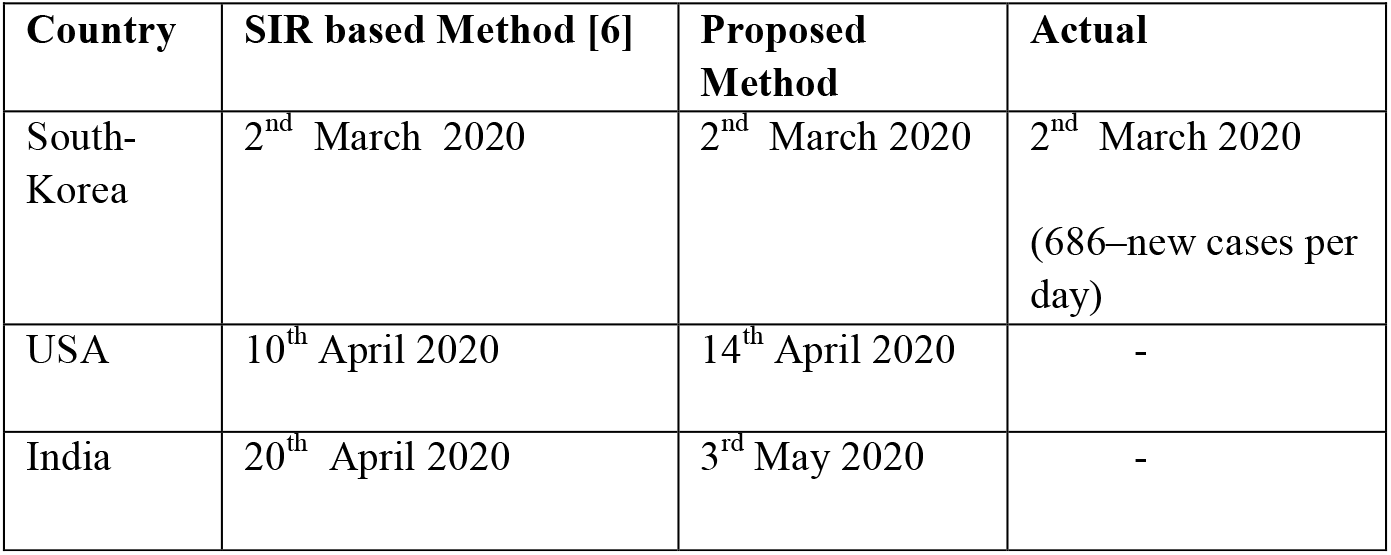
Comparison of the results for the peak position.

**Table 2:** Comparison of the results for the ending life-cycle.

| Country |  | SIR based Method [6] | Proposed Method | Actual |
| --- | --- | --- | --- | --- |
| South-Korea | 97% | 22<sup>nd</sup> March 2020 | 11<sup>th</sup> March 2020<br>(Accumulated Cases-7597 ) | 11<sup>th</sup> March 2020<br>(Accumulated Cases-7755 ) |
|  | 99% | 31<sup>st</sup> March 2020 | 13<sup>th</sup> March 2020<br>(Accumulated Cases-7709 ) | 13<sup>th</sup> March 2020<br>(Accumulated Cases-7979 ) |
| USA | 97% | 12<sup>th</sup> May 2020 | 12<sup>th</sup> May 2020 | - |
|  | 99% | 24<sup>th</sup> May 2020 | 18<sup>th</sup> May 2020 | - |
| India | 97% | 22<sup>nd</sup> May | 5<sup>th</sup> June 2020 | - |
|  | 99% | 1<sup>st</sup> June | 13<sup>th</sup> June 2020 | - |

Tables-1 and Table-2 show that our simulation results for the detection of the peak position and estimating ending life-cycle of the COVID-19 curve for South–Korea are very close to the actual values, known to us. The actual time-series (NCPD) of South-Korea, also confirms the peak on the 2^nd^ March 2020, with a value of 686 new cases per day. Our simulation results for South-Korea, also suggested that 97 % & 99 % of the total affected cases will be reported by the 11^th^ and 13^th^ of March 2020 with accumulated cases values of 7597 and 7709 respectively. Actual time-series (NCPD) of the South-Korea reveals that on these days, 7757 and 7979 accumulated cases were reported respectively.

We have not mentioned the actual percentage for these values because our method detect only the major wave. While comparing our simulation results to the SIR based method [6], the peak position days for South Korea and the USA are observed to be very close but peak position day for India differs. More data points in the future may clarify the reason for this difference. The same reason also applies to the differences in 97% and 99% ending values of the COVID-19 curve for India. While estimating the ending life-cycle, we assume that the fall of the curve from its peak position will follow an ideal pattern similar to that of South-Korea, but practically this may not happen for other countries. So, a minor modification in the ending life-cycle values may be made based on the dynamics of the relative position of the peak. We also investigated the variation in values of the coefficients *a, b* and *c* of the eqn. (1) for South-Korea and the USA. The variation in the values of these coefficients indicates the changing dynamics of the COVID-19 curve and estimating these coefficients in advance may also prove useful. A good perspective regarding predictive monitoring for the COVID-19 curve and its interpretation is presented in [6]. We also recommend the reading of these interpretations so as not to misunderstand the predictive monitoring.

## 4. Conclusion

Detection of the flattening of the COVID-19 curve and estimating its ending life-cycle is useful to contain the spread of the disease through appropriate policy interventions. A novel method for detection of flattening of the COVID-19 curve and estimating its ending life-cycle using only the time-series of new cases per day, grossly fitted to the Gaussian function is proposed. Simulation results using the COVID-19 curve for the South-Korea demonstrate the ability of the method to detect the peak as well as estimating its ending life-cycle. Further, comparisons of the simulations results to the method based on the SIR model for the USA and India, suggest that the proposed method can quantify the varying dynamics of the COVID-19 curve without utilizing critical epidemiological parameters.

## Data Availability

Data has been obtained from our world in data website

https://ourworldindata.org/coronavirus-data

## Notes

### Competing Interest Statement

The authors have declared no competing interest.

### Clinical Trial

This is not a clinical trial

### Funding Statement

No funding has been received

## References

1. Li. Q. et al. Early Transmission Dynamics in Wuhan, China, of Novel Coronavirus-166 Infected Pneumonia. N Engl J Med, doi:10.1056/NEJMoa2001316 (2020)

2. N. M. Ferguson et al.,. Impact of non-pharmaceutical interventions (npis) to reduce covid-19 mortality and healthcare demand. London: Imperial College COVID 19 Response Team, March 16 (2020), 10.25561/77482.

3. Andrea Remuzzi, Giuseppe Remuzzi. COVID-19 and Italy: what next? the Lancent 2020; https://doi.org/10.1016/S0140-6736(20)30627-9

4. https://www.ecdc.europa.eu/en/covid-19-pandemic

5. https://coronavirus.jhu.edu/map.html

6. J. Luo, Predictive Monitoring of COVID-19. https://ddi.sutd.edu.sg/

7. S. Woody et al. Projections for first-wave COVID-19 deaths across the US using social-distancing measures derived from mobile phones, https://www.medrxiv.org/content/10.1101/2020.04.16.20068163v2

8. https://ourworldindata.org/coronavirus-source-data

9. https://www.who.int/emergencies/diseases/novel-coronavirus-2019

